# Leveraging a genetic proxy to investigate the effects of lifelong cardiac sodium channel blockade

**DOI:** 10.1101/2025.04.15.25325909

**Authors:** Julian S. Wanner, Maren Krafft, Teemu Niiranen, Dominic S. Zimmerman, FinnGen, Patrick Ellinor, Girish Nadkarni, Sean J Jurgens, Joel Rämö, Henrike O. Heyne

## Abstract

Atrial fibrillation (AFib) and other cardiac arrhythmias pose a major public health burden but prevention remains difficult. Here, we investigated a genetic variant which we found to act like a natural lifelong cardiac sodium channel blockade.

We studied the impact of the Finnish-enriched *SCN5A* missense variant (rs45620037 [T220I]) on cardiac arrhythmias, associated mortality and electrocardiographic (ECG) phenotypes in > 1M individuals across three cohorts (FinnGen, UK biobank, Health 2000). We identified protective effects of T220I on multiple common cardiac arrhythmias, most notably atrial fibrillation (AFib) (hazard ratio [HR] 0.60, 95% confidence interval [CI] 0.55 – 0.66, p = 3.13×10^-25^), but also ventricular premature depolarization or ventricular tachycardia, yet increasing susceptibility to conduction-slowing conditions such as sick sinus syndrome (mostly in older age groups). Overall, T220I conveyed a protection from mortality due to cardiac arrhythmia (HR = 0.65, 0.47 – 0.98, p = 0.015) without a significant effect on overall mortality (HR=0.98, p = 0.78). T220I heterozygotes had similar electrophysiological effects as sodium channel blockers such as significantly shortening QT intervals (−7.49 ms 95% CI −10.07 – [−4.91] ms, p = 0.0037, n=6,048) in the H2000 cohort, which we replicated in the UK Biobank (n=81,195). In addition, T220I protected from (left) heart failure and dilated cardiomyopathy. Early post–myocardial infarction, we found that T220I increased mortality in agreement with known sodium channel blocker effects which however continuously normalised to baseline 10-15 years post myocardial infarction. Finally, we found that T220I could lower a high genetic burden (a high polygenic score) for AFib to population average.

The *SCN5A* T220I variant, consistent with a previously described weak loss-of-function effect, acted like a genetic proxy for cardiac sodium channel blockade. This enabled us to gain new potentially clinically relevant insights for pharmacological sodium channel blockade such as after myocardial infarction which would be too risky to investigate with clinical trials. Our findings may also inspire redesign of cardiac sodium channel blockers.

## Introduction

Cardiac arrhythmias pose a major health burden. Atrial fibrillation (AFib) is the most common type with over 4 million incident cases and 300,000 deaths per year globally ^1^. It increases the risk for many cardiovascular outcomes including heart failure and stroke and is associated with increased all-cause mortality. However, prevention and control of atrial fibrillation and its sequelae remains difficult, partially due to heterogeneous causal mechanisms. Both common and rare genetic factors have been previously described to have large influences on the development of cardiac arrhythmias. Specifically, AFib has been shown to be highly polygenic ^2^.

Ion channels are essential for cardiac signaling. The *SCN5A* gene encodes Na_V_1.5, the pore-forming subunit of the cardiac sodium channel responsible for the initiation and propagation of cardiac action potentials. Thus, genetic variants within *SCN5A* can increase the risk for certain cardiac arrhythmias and arrhythmia syndromes, such as Brugada syndrome (BrS)^3,4^ and AFib ^2,5–7^.

The different types of clinical presentations caused by *SCN5A* mutations can largely be explained by the degree, and specific type of sodium channel dysfunction, at the molecular level. Loss-of-function (LoF) of Na_V_1.5 (such as caused by protein truncating variants) are a main cause of Brugada syndrome (BrS) ^3^, as well as conduction deficits resulting in bradycardia, conduction blocks, or sick sinus syndrome (SSS). In contrast, gain of channel function (GoF) has been linked to other specific disease entities, such as long QT syndrome ^3,8,9^. More severe phenotypes such as congenital SSS can be caused by more severe LoF of the channel, for instance through the presence of two defective *SCN5A* alleles. ^10–12^ Mechanistically, genetically caused *SCN5A* LoF is comparable to sodium channel blocker medications, which are used to mediate a desirable LoF effect, but may also increase risk of cardiac arrhythmias in certain contexts.

In a recent study of 180k Finnish individuals ^13^, we discovered that carriers of one genetic missense variant in SCN5A, T220I (rs45620037 or NM_000335.5[SCN5A]:c.659C>T [p.T220I]) had only half the risk of AFib compared to the general population ^14^. The T220I variant is 5-fold enriched in the Finnish population to an allele frequency of 0.46%, compared to a frequency of 0.11% in the UKBB and 0.086% in non-finnish European populations^15^. The clinical significance of T220I remains uncertain based on limited data from family studies and case reports (for an overview please see Supplementary Table 1). T220I was previously described in individuals with childhood-onset SSS who also carried a LoF variant on the other allele^16^, however, the evidence was confined to a few families ^17–19^. Due to the high population frequency and absence of large effects on disease, it was classified as a likely benign variant (5 independent submissions) or variant of uncertain significance (4 independent submissions) in ClinVar, a public variant database ^20,21^. The T220I variant was also associated with AFib^11^ and dilated cardiomyopathy ^11,22,23^ in individual case reports. An association of T220I with BrS could not be established using a Flecainide test^24^, which uses the class Ic sodium channel blocker Flecainide to reveal BrS-specific ECG patterns^25^. Electrophysiological studies showed a weak LoF ^17,18^, due to a stabilized inactivation state and thus reduced sodium current, the sinus node’s ability to reach the threshold for action potential firing is reduced, leading to reduced electrical signaling within the heart and thus potential bradyarrhythmic effects ^26,27^. T220I’s effects on slowing conduction are in line with a decreased risk for diseases associated with reentry such as atrial fibrillation, in line with our earlier observations ^13^.

Due to the previously established mild LoF effect we hypothesized that T220I acts as a proxy for a mild sodium channel blocker. We thus further hypothesized that this should affect cardiac conduction reflected in specific ECG changes. In addition, T220I should affect risk for specific cardiac diseases such as protection from AFib (we observed earlier) but also risk-increasing effects for sinus bradycardias as a common side effect of cardiac sodium channel blockers and as the variant was earlier associated with SSS ^19,20^. In addition, we hypothesized a potential increased risk for other possible side effects of cardiac sodium channel blockers in T220I carriers such as conduction blocks, ventricular fibrillation, Brugada syndrome–like changes or Torsades de pointes.

To investigate the effects of T220 on protection and risk from different cardiac phenotypes including mortality and conduction parameters using ECG data we leveraged data from three mostly population-based research cohorts – FinnGen^14^ , the UK Biobank^28^ (UKBB), and the Finnish cohort Health 2000 (H2000)^29^ - using data from a total of 1,014,635 individuals. The primary analysis was conducted within FinnGen including longitudinal electronic health record data covering up to five decades in over 520,000 individuals ^14^ a cohort of almost 3 times the size compared to when we first discovered the variant’s protective effect ^13^.

## Methods

### Study Design and Cohorts

We conducted this study using three primary cohorts: FinnGen, UKBB, and H2000. Both the UKBB and H2000 are population-based research cohorts with genetic and phenotypic data, initiated in 2006 and 2000, respectively.The FinnGen cohort consisted of 520,210 individuals with longitudinal health and genetic data, across 70 years including genotype and phenotype information from national health registers. Further details about the cohort have been previously described^14^, while case numbers for specific cardiac arrhythmias are shown in Table 2. The UK Biobank is a prospective study cohort of 500,000 participants from the United Kingdom aged between 40 and 69 at recruitment. Automated ECG measurements (PR interval, P-wave duration, QRS duration, and QT interval) were available for 116,015 UK Biobank study participants, extracted either from 12-lead ECGs (N = 67,621 with at least one measurement released centrally by UK Biobank) or 3-lead upright resting ECGs taken before a bicycle exercise test (N = 60,755 with at least one measurement extracted using the GE Marquette 12SL ECG analysis program). We evaluated two sets of measurements: 1) measurements derived only from 12-lead ECGs and 2) measurements derived from either 12-lead or 3-lead ECGs, prioritizing 12-lead ECGs in participants who had measurements from both 12-lead and 3-lead ECGs (N = 12,361). The H2000 cohort comprised 6,048 participants with ECG measurements^29^ and included a health examination interview, a physical examination, and an extensive questionnaire.

**Table 1.** Descriptive statistics of the participating biobanks. Sample sizes, median age at recruitment with interquartile range (IQR), median follow-up duration with IQR, proportion of female participants, and ascertainment strategies.

| Study | Sample size | Age of recruitment (yrs)<br>median (IQR) | %Female | Ascertainment<br>Strategy |
| --- | --- | --- | --- | --- |
| FinnGen | 520,210 | 55.5 (41.7 – 69.2) | 56.4 | Population & Hospital |
| H2000 | 3,884 | 47.7 (35.5 – 59.9) | 51.9 | Population-based |
| UK Biobank | 108,186 | 57.00 (50.00 – 62.00) | 51.6 | Population-based |

**Table 2.** Cases and Controls for ICD codes and Genotypes

| ICD Code | Description | Non-carrier |  | Heterozygous |  |
| --- | --- | --- | --- | --- | --- |
|  |  | Cases | Controls | Cases | Controls |
| I42.0 | Dilated cardiomyopathy | 4548<br>(0.9%) | 510426 | 26<br>(0.5%) | 5077 |
| I44.0 | First degree atrioventricular block | 1555<br>(0.3%) | 513419 | 20<br>(0.4%) | 5083 |
| I44.2 | Third degree atrioventricular block | 3231<br>(0.6%) | 511743 | 39<br>(0.8%) | 5064 |
| I44.7 | Other/unspecified atrioventricular block | 2278<br>(0.4%) | 512696 | 9<br>(0.2%) | 5094 |
| I45.2 | Bifascicular block | 481<br>(0.1%) | 514493 | 6<br>(0.1%) | 5097 |
| I45.9 | Unspecified conduction disorder | 1148<br>(0.2%) | 513826 | 11<br>(0.2%) | 5092 |
| I47.1 | Paroxysmal supraventricular tachycardia | 8561<br>(1.7%) | 506413 | 46<br>(0.9%) | 5057 |
| I47.2 | Ventricular tachycardia | 3266<br>(0.6%) | 511708 | 16<br>(0.3%) | 5087 |
| I48.0 | Paroxysmal atrial fibrillation | 10466<br>(1.4%) | 504508 | 51<br>(1%) | 5052 |
| I48.1 | Persistent atrial fibrillation | 3137<br>(1.1%) | 511837 | 15<br>(0.3%) | 5088 |
| I48.2 | Chronic atrial fibrillation | 2878<br>(0.6%) | 512096 | 10<br>(0.2%) | 5093 |
| I48.9 | Atrial fibrillation, unspecified | 7101<br>(1.4%) | 507873 | 32<br>(0.6%) | 5071 |
| I48 | Atrial fibrillation and flutter | 48314<br>(9.3%) | 466660 | 287<br>(5.6%) | 4816 |
| I49.1 | Premature atrial contractions | 6061<br>(1.1%) | 508913 | 40<br>(0.8%) | 5063 |
| I49.3 | Paroxysmal tachycardia, unspecified | 14814<br>(2.8%) | 500160 | 75<br>(1.4%) | 5028 |
| I49.4 | Other and unspecified cardiac arrhythmias | 7022<br>(1.3%) | 507952 | 43<br>(0.8%) | 5060 |
| I49.5 | Sick sinus syndrome | 5089<br>(1%) | 509885 | 109<br>(2.1%) | 4994 |
| I49.9 | Cardiac arrhythmia, unspecified | 28906<br>(5.6%) | 486068 | 211<br>(3.4%) | 4892 |
| I50.1 | Left ventricular failure | 7136<br>(1.4%) | 507838 | 55<br>(1.1%) | 5048 |
| I50.9 | Heart failure, unspecified | 20451<br>(3.9%) | 494523 | 174<br>(3.4%) | 4929 |
| I50 | Heart failure | 2847<br>(0.6%) | 512127 | 21<br>(0.4%) | 5082 |

Collected samples include venous blood samples, measurements of height, weight, and blood pressure, psychometric tests, and a resting 12-lead ECG, amongst others. The clinical examination included the medical history, an assessment of drug prescriptions, the cardiovascular and pulmonary status. The ECG was automatically interpreted by the Social Insurance Institution’s Research Center. We obtained ethical approvals from the respective institutional review boards, and all participants provided informed consent for biobank-based research.

### Phenotype Definitions

We identified cardiac arrhythmias, including atrial fibrillation (AFib) and Brugada syndrome (BrS), using ICD codes I40–I50 and their subcategories (I40.0–I50.9) from national healthcare registries in FinnGen, while mortality outcomes were derived from national death registries, with a total of 1.87 million cardiac arrhythmia cases and 9,771 deaths due to arrhythmia reported. We defined cases as individuals with respective diagnosis codes and did not exclude older individuals to capture potential long-term protective effects. Individuals missing required clinical or genotyping data were removed.

From the H2000 study, we initially included 6,048 individuals, of whom 62 were heterozygous for T220I. To control for potential confounders, we included covariates such as age, sex, BMI, medication use (including psychotropic drugs, antidepressants, and glaucoma medication), diseases, and other factors affecting heart function. We performed sensitivity analyses and excluded individuals with outlier or missing covariate values as follows.

We applied the following exclusion criteria: individuals older than 90 years, those in poor health with a fitness index below 54, and individuals classified as morbidly obese with a BMI over 40. We also excluded individuals with abnormal blood test results (calcium concentration <2.2 or >2.6 mmol/l, or TSH >4.5 mU/l), extreme blood pressure (diastolic >120 mm Hg or systolic >180 mm Hg), and those taking heart medications, including cardiac glycosides, antiarrhythmics, beta blockers, calcium antagonists, and thrombosis prevention drugs. After excluding 1,555 wild-types and 18 heterozygotes taking heart medications, we reduced the cohort to 3,884 individuals.

For downstream ECG analyses using Framingham correction, we further restricted the dataset to individuals with a heart rate between 60–100 bpm, resulting in a final dataset of 3,188 individuals.

We excluded participants with pacemakers (based on automated ECG reports or health record data), Wolff-Parkinson-White syndrome, class I or III antiarrhythmic use, or digoxin use. For analyses of the corrected QT interval, we conducted analyses both before and after excluding participants with extreme heart rates (<40 or > 120 beats per minute) or QRS interval >120ms. Heart rate–corrected QT intervals were calculated using the Bazett formula, defined as QTc = QT(ms)/√RR(s).

Details of genotyping and initial quality control have been described previously ^28^. The *SCN5A* variant T220I was directly genotyped. Prior to genomic analyses, we excluded participants whose submitted gender did not match their gender inferred from genotypes, participants who were outliers in heterozygosity or missingness, participants with putative sex chromosome aneuploidy, and participants with >5% genotype missingness rate. We restricted the analyses to the white British ancestry subset and removed related participants based on a kinship threshold of 0.0884.

### Cardiac Arrhythmia Association Analysis

We performed an association study between the T220I variant and case/control status for ICD categories I40–I50 and their subcategories using the PheWAS R package (v0.99.6-1). To detect potential non-additive genetic effects, we analyzed heterozygous and homozygous carriers separately. To correct for multiple testing across 148 tests, we applied a Bonferroni threshold of 3.37 x 10^-4^.

For subsequent survival analysis, we used Cox proportional hazard models implemented in the R package survival (v3.7.0). Follow-up time began at birth and ended at the age of first diagnosis for cases or at the age of the last recorded entry or death, whichever occurred first. We adjusted the models for the first ten principal components of genetic ancestry, sex and genotyping batch to account for technical artifacts. To ensure adequate power to detect both protective and risk-enhancing effects of T220I, we performed power calculations with a power of 0.8 and an alpha of 0.05 using the R package powerSurvEpi (v0.1.1.3).

All statistical tests were performed as two-sided, without assuming a specific direction of effect.

### Sensitivity Analyses

We conducted sensitivity analyses for our medication analysis using linear mixed models (R v4.2.4) to adjust for potential confounders, including medication use and age at diagnosis. We filtered drug reimbursements from the KELA register in FinnGen using ATC codes C01BC04, C07AB03, C07AB07, C07AG02, C07AB02, C07AA07, C01BD01, C01BD07, C08DB01, C08DA01, and C01AA05, totaling 7.05 million drug reimbursements.

### PGS Calculation and Prediction

We estimated PGSs for AFib and BrS using PRS-CS ^38^, a Bayesian method that incorporates linkage disequilibrium. To construct disease-specific PGSs, we used publicly available GWAS summary statistics for AFib ^39^ and BrS ^4^ and standardized scores to a mean of 0 and a standard deviation of 1. To avoid potential LD effects, not captured by PRS-CS, we excluded genetic variants located ±500 kb around T220I.

To assess whether the protective effect of T220I was additive with an AFib PGS, we performed an interaction analysis with the survival package between T220I carrier status and the PGS, treating the PGS as a continuous variable.

We further categorized individuals into disease-specific risk bins: high PGS (>1 SD above the mean), low PGS (< −1 SD below the mean) and average PGS (between −1 SD and 1 SD) and conducted survival analysis across the lifetime using our usual covariates in addition to PGS bins. The PRS-CS pipeline used in FinnGen is detailed at https://github.com/FINNGEN/CS-PRSpipeline.

### Genomic association analyses of electrocardiographic data in the UK Biobank and H2000

We evaluated the association of T220I with each of four electrocardiographic measurements (QRS, PR, QT, P-dur) using linear regression as implemented in the glm function in R (version 4.3.0), with genotype-inferred sex, age during the ECG visit, the first 10 genomic principal components, and the genotyping array as covariates. Correction was done according to Bazett.

Within H2000, we conducted multiple linear regression analyses with Framingham-corrected QTc as the outcome variable. Independent variables included genotype, sex, age, and interaction terms (age*sex, age*age). We further adjusted for covariates such as calcium, TSH, systolic and diastolic blood pressure, BMI, fitness index, illnesses (respiratory, arrhythmia, and heart disease), medication use, and the first four principal components of genetic ancestry.

## Results

### Description of cohorts

We set out to test the effects of T220I on lifetime risk of cardiac diseases using three different cohorts: FinnGen, the UK Biobank, and H2000. We used FinnGen (data freeze 12) for our main analysis, which included 520,210 individuals. In FinnGen, the allele frequency of T220I was 0.005, with 28 homozygotes and 5,021 heterozygotes. Further descriptive statistics of the three cohorts are shown in Table 1.

### The effect of T220I on common cardiac arrhythmias

As T220I was previously found to cause mild LoF of the Na_V_1.5 channel, it may be a genetic proxy for a lifelong mild sodium channel blockade. We thus explored the effect of T220I on diseases associated with sodium channel blockade. Specifically, we tested whether heterozygotes of T220I are associated with cardiac arrhythmias and heart failures, testing cardiac-related ICD codes I40 – I50 within FinnGen.

We found that in a heterozygous state, T220I was associated with protection from multiple types of tachycardia such as AFib (odds ratios [OR] 0.58, 95% CI 0.47 – 0.69, p = 8.34×10^-22^), which we replicated in the UKBB (see Supplementary Note 1), or ventricular premature depolarization (see Figure 1a). However, it also increased risk of cardiac bradyarrhythmias, such as SSS (OR 2.36, 95% CI 2.19 – 2.53, p = 4.32×10^-22^) (Figure 1b and c). Disease associations were relatively unaffected by the removal of related individuals. Cases and controls are shown in Table 2. We also found a protective effect of T220I heterozygosity on ventricular tachycardia (OR = 0.61, 95% CI 0.26 – 0.98, p = 5.7×10^-3^) we did not necessarily expect considering known side effects of long-term sodium channel blocker used for prevention of AFib. Further sensitivity analyses (method: linear mixed model) revealed that the protective effects were not due to an earlier age of disease onset or medications (see Supplementary Note 2). In homozygotes, we found an increased risk for bradyarrhythmias, specifically bifascicular block (OR = 7.88, 95% CI 7.11 – 8.66, p = 1.76×10^-07^) and sick sinus syndrome (OR = 3.81, 95% CI 3.27 – 4.36, p = 1.27×10^-06^).

**Figure 1.**
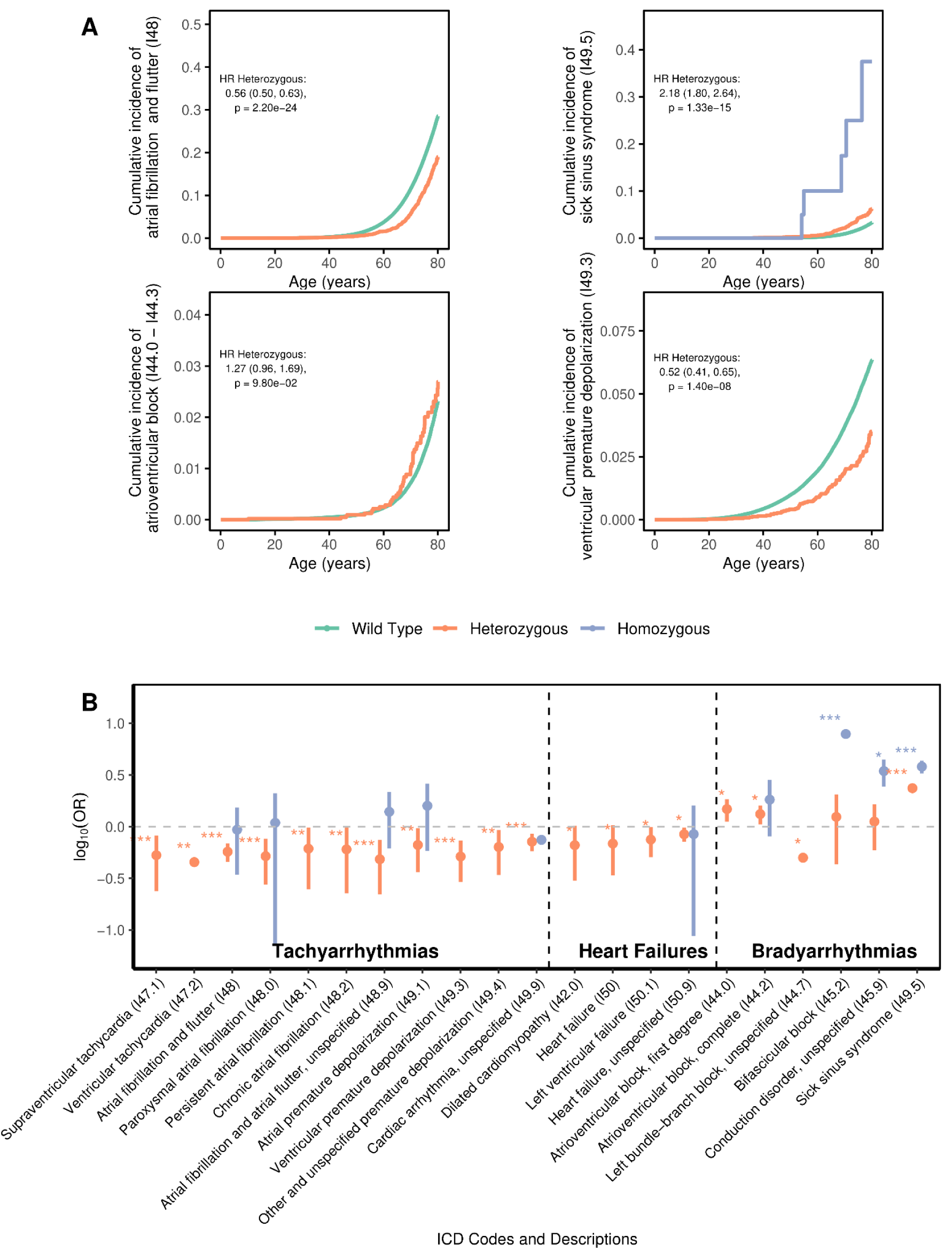
Effect of the SCN5A variant T220I on specific cardiac arrhythmias. (a) Age at first diagnosis (x-axis) of different cardiac arrhythmias, separately for different T220I carrier status. Cumulative disease incidence is given on the y-axis, hazard ratios of T220I effect across lifetime with 95% confidence intervals and respective p-values are shown in the figures. T220I homozygotes are colored in blue, T220I heterozygotes in orange and wild types in green. (b) log_10_ of odds ratios (OR) of significant associations of T220I with cardiac phenotypes I40 to I50. Statistical significance prior to multiple testing correction (method: Bonferroni) is denoted by asterisks: * = p < 0.05, ** = p < 0.01, *** = p < 0.001. Homozygous carrier states are colored in blue, heterozygotes in orange.

Next, we explored the effects of T220I on cardiac diseases across the lifetime. Most effects remained consistent across the life course (Figure 1a). Heterozygotes were protected against cardiac arrhythmias throughout their lifetime, but were also at an increased risk of bradyarrhythmias, such as SSS and AV block; survival curves are shown in Figure 1a. Consistent with this, we found a higher rate of pacemaker procedures in T220I heterozygotes (HR = 1.50, 95% CI 1.31 – 1.72, p = 5.14×10^-09^) and homozygotes (HR = 4.12, 95% CI 1.54 – 11.00, p = 4.73×10^-03^); see Supplementary Figure 1.

Exploring the effect of T220I on heart failure and cardiomyopathies we found that T220I nominally protected against dilated cardiomyopathy (HR = 0.66, 95% CI 0.46– 0.94, p = 0.021) and heart failure (HR = 0.84, 95% CI 0.75– 0.93, p = 1.24×10^-03^) across the lifetime. To test whether we observed fewer protective effects in homozygotes due to lack of power - or due to lack of effect - we conducted a power analysis (R package: powerSurvEpi (v0.1.3)), which indicated that currently, we did not have the necessary sample size to observe protective effects in individuals with AFib (n=28, n_required_=148).

### T220I protects from death due to cardiac arrhythmias

As T220I decreased but also increased risk for multiple cardiac arrhythmias, we wanted to explore whether T220I had a net positive or negative overall effect on death due to cardiac arrhythmias across the lifetime. Overall, we found a protective effect from mortality due to cardiac arrhythmias (HR = 0.65, 95% CI 0.46 – 0.92, p = 0.015); see Figure 2. We found no effect on overall cardiovascular (HR=0.99, 95% CI 0.85 – 1.15, p = 0.90) or all-cause mortality (HR=0.98, 95% CI 0.88 – 1.09, p = 0.78); however, this was expected since we found no effect of T220I on any diseases not originating in cardiac cells (exploratory analysis, see Supplementary Figure 3). After adjusting for competing risks due to other causes of death we still observed T220I’s protective effect on death due to cardiac arrhythmias; see Supplementary Figure 4. We think this effect was mainly driven by T220I’s protective effect against deaths due to AFib across the lifetime (HR 0.58, 95% CI 0.37 – 0.85, p = 6.85×10^-03^).

**Figure 2.**
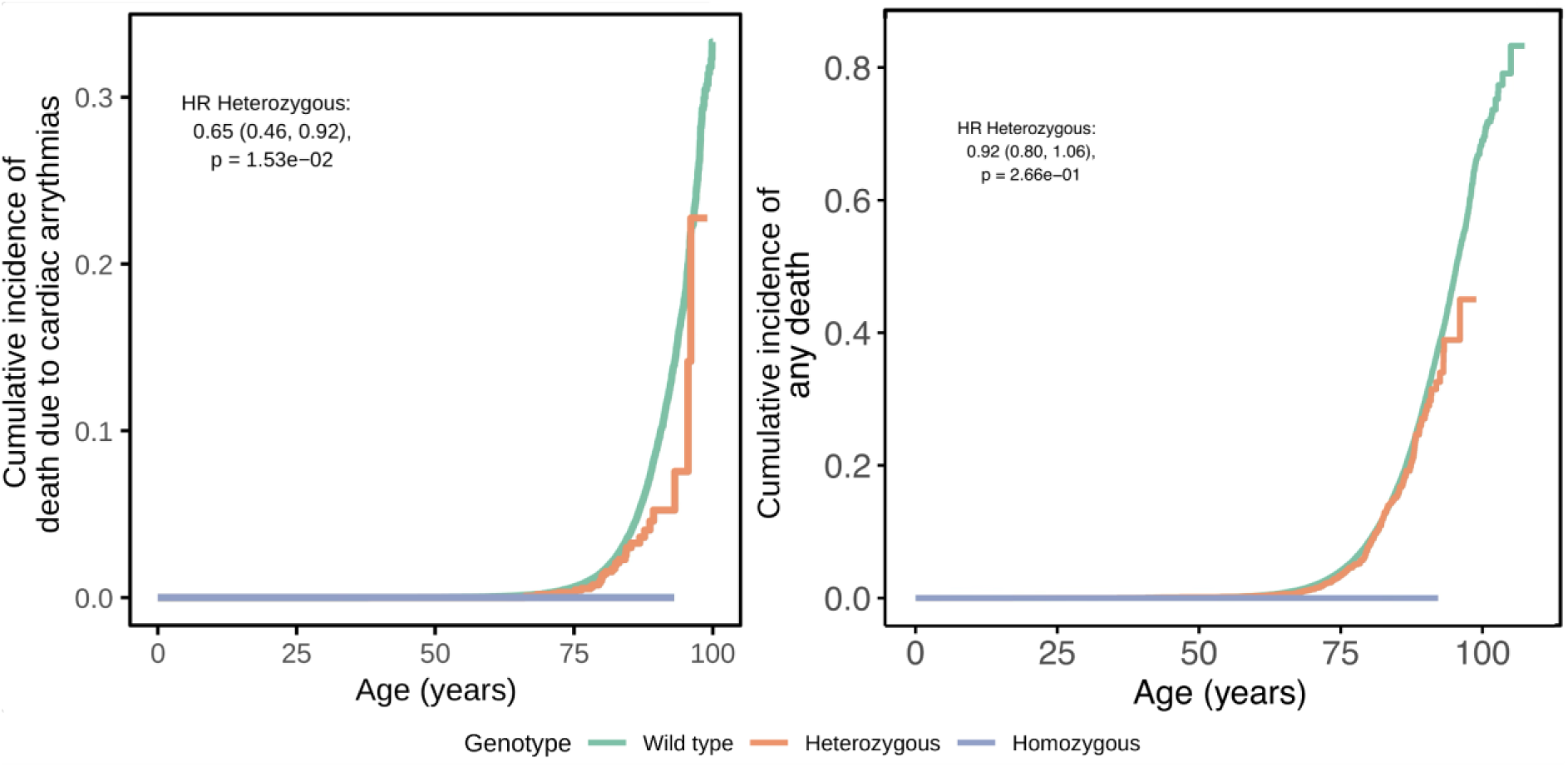
T220I protects from death due to cardiac arrhythmias with no effect on all-cause mortality. (a) Death due to cardiac arrhythmias (b) Death due to any cause. Data are shown as Kaplan-Meier survival estimates, significant hazard ratios across the lifetime are shown inside the survival plots. Heterozygotes are colored in orange, wild types in green and homozygotes in blue.

### Associations of T220I with outcomes after myocardial infarction

We next sought to investigate the effect of T220I in individuals with structural heart disease, motivated by the finding of increased mortality in patients who received sodium channel blockers after a myocardial infarction in the CAST trial ^30^. In 25,307 individuals who had suffered a myocardial infarction, we found that 248 T220I carriers were still protected from AFib (HR = 0.66, 95% CI 0.48 – 0.89, p = 0.007). After myocardial infarction, T220I’s protective effect from cardiac arrhythmia related mortality stayed similar, although it was not significant due to the reduced sample size (HR 0.44, 95% CI 0.18 – 1.06, p = 0.067), with no effects on all-cause mortality (HR 1.12, 95% CI 0.95 – 1.31, p = 0.18) or cardiovascular mortality (HR 1.06, 95% CI 0.95– 1.19, p = 0.3). However, when restricting our observation period to 10 months post–myocardial infarction to simulate the CAST trial, we also found an increased all-cause mortality in the group of T220I carriers, similar to pharmacologic sodium channel blockade, up to 3 years after a myocardial infarction. Shifting our observation windows to individuals surviving longer after myocardial infarction, we found however that T220I associated mortality continuously decreased reaching baseline 15 years post–myocardial infarction; see Supplementary Figure 5 and Supplementary Note 3.

### T220I alters the effects of common genetic variation on AFib

Next, we wanted to test how T220I influences the effect of common genetic variation on AFib. Thus, we computed a disease-specific PGS for AFib (method: PRS-CS^1^) and tested its association with AFib risk in relation to T220I carrier status in FinnGen. As expected, a higher AFib-PGS increased risk for AFib across the lifetime (HR per SD = 1.66, 95% CI 1.65 – 1.67, p < 2.23×10^-308^). However, T220I could counterbalance the effect of a high AFib-PGS on AFib risk. As an example, individuals with a high AFib-PGS (PGS > +1 SD) who also carried T220I had an average risk for AFib across the lifetime (Figure 3). This association remained significant after the removal of all variants within ±500 Kb of T220I from the PGS to exclude potential variants that could capture the effect of T220I due to linkage disequilibrium. Surprisingly, we found that the variant significantly modulated how common genetic variation affects AFib risk (attenuation factor: 0.88, 95% CI 0.81 – 0.99, interaction p = 0.017, PGS - HR per SD in variant carriers: 1.50, 95% CI 1.36 – 1.67, p = 2.95×10^-15^)), which we could not attribute to sex or age interactions (Supplementary Figure 2a and b).

**Figure 3.**
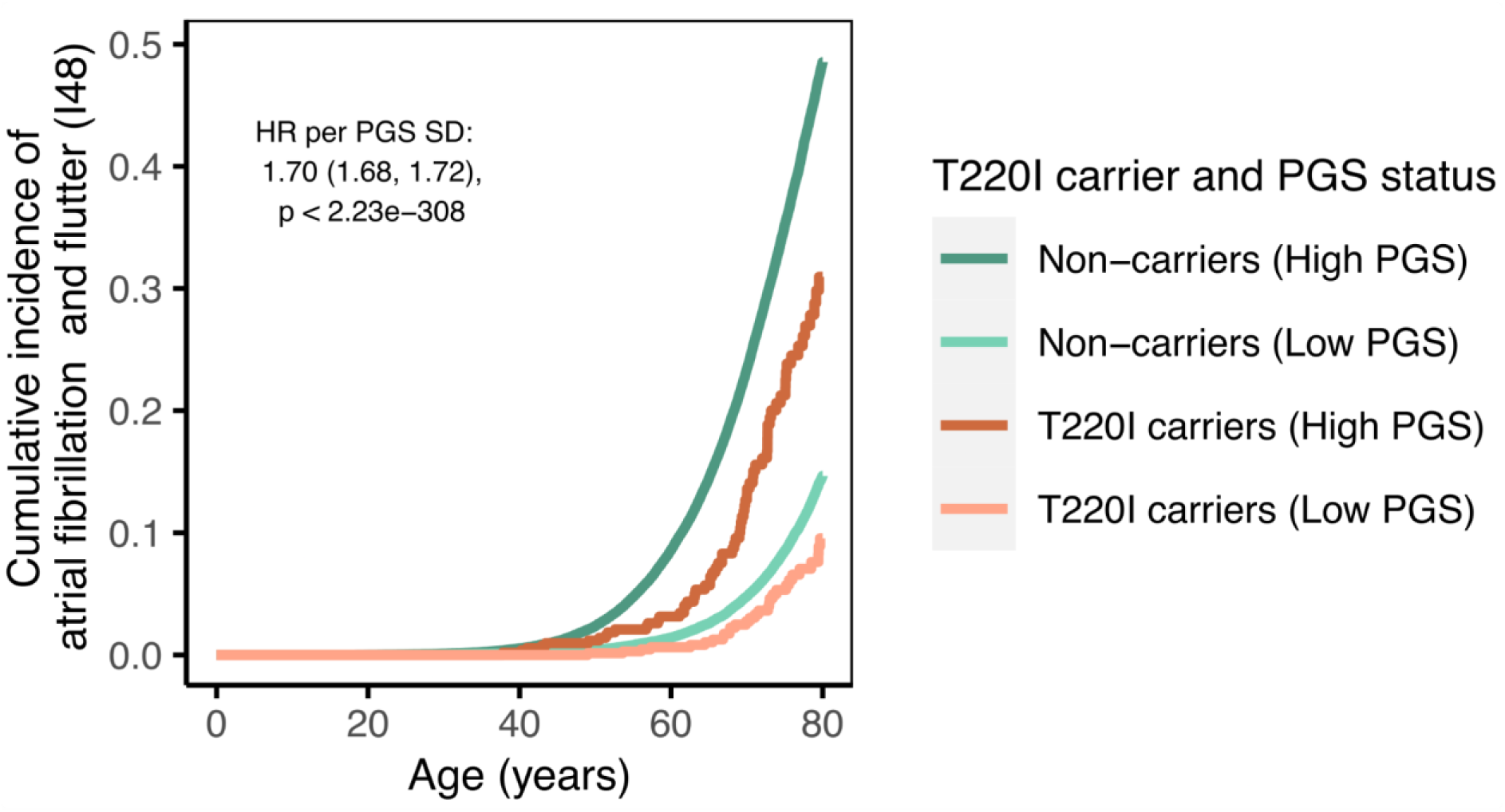
T220I lowers a high genetic risk for AFib. Age at first diagnosis of AFib, stratified by carrier status of T220I and high or low AFib-PGS. Data are shown as cumulative incidence of AFib (y-axis) across the lifetime (age in years, x-axis). Hazard ratio (HR) of AFib-PGS effect across lifetime is shown inside the Figure. Heterozygotes of T220I are colored in orange shades and wild types in green shades. Low PGS group defines individuals with an AFib-PGS < −1 SD, “High PGS” indicates individuals with an AFib-PGS > +1 SD.

### T220I is associated with shortened QT and PR intervals

Lastly, we evaluated how the T220I variant affects cardiac conduction investigating five ECG parameters. We found a significantly shortened QT intervals (QTc, Bazett corrected) in the H2000 cohort (−7.49 ms, 95% CI −10.07 ms – [-4.91] ms, p = 0.0037, method: linear regression. n=3188) and in the UK Biobank after the exclusion of participants with prolonged QRS duration or extreme heart rates (−4.94 ms, 95% CI −8.54 – [-1.32] ms, p = 7.36×10^-03^, n=82242) (Figure 4). Effect sizes of T220I on QTc time remained similar in a smaller dataset including only 12-lead ECGs in the UK Biobank (−4.65 ms, 95% CI −9.94 – [-0.64] ms, p = 0.08). The associations also remained significant when using a different QT correction method (Framingham) or adjusting for effects of medications, fitness parameters and blood pressure (see sensitivity analyses in Supplementary Note 4). We also found associations with PR time in both the 12-lead (11.04 ms 95% CI 5.01, 17.07 p = 3.30×10^-4^) and the combined 12-lead and 3-lead ECG datasets (5.74 ms 95% CI 1.809, 9.68, p = 0.0042 in the UK Biobank (Supplementary table 2). In H2000, we found no effect on PR time. In addition, we searched for abnormalities in real ECGs of T220I carriers in the H2000 cohort (n=62) and the UK Biobank (n=65). We found no Brugada-like patterns in the H2000 cohort and the UK Biobank, but a high number of sinus bradycardia in the H2000 cohort, in line with increased bradycardia associated diseases in variant carriers in FinnGen.

**Figure 4.**
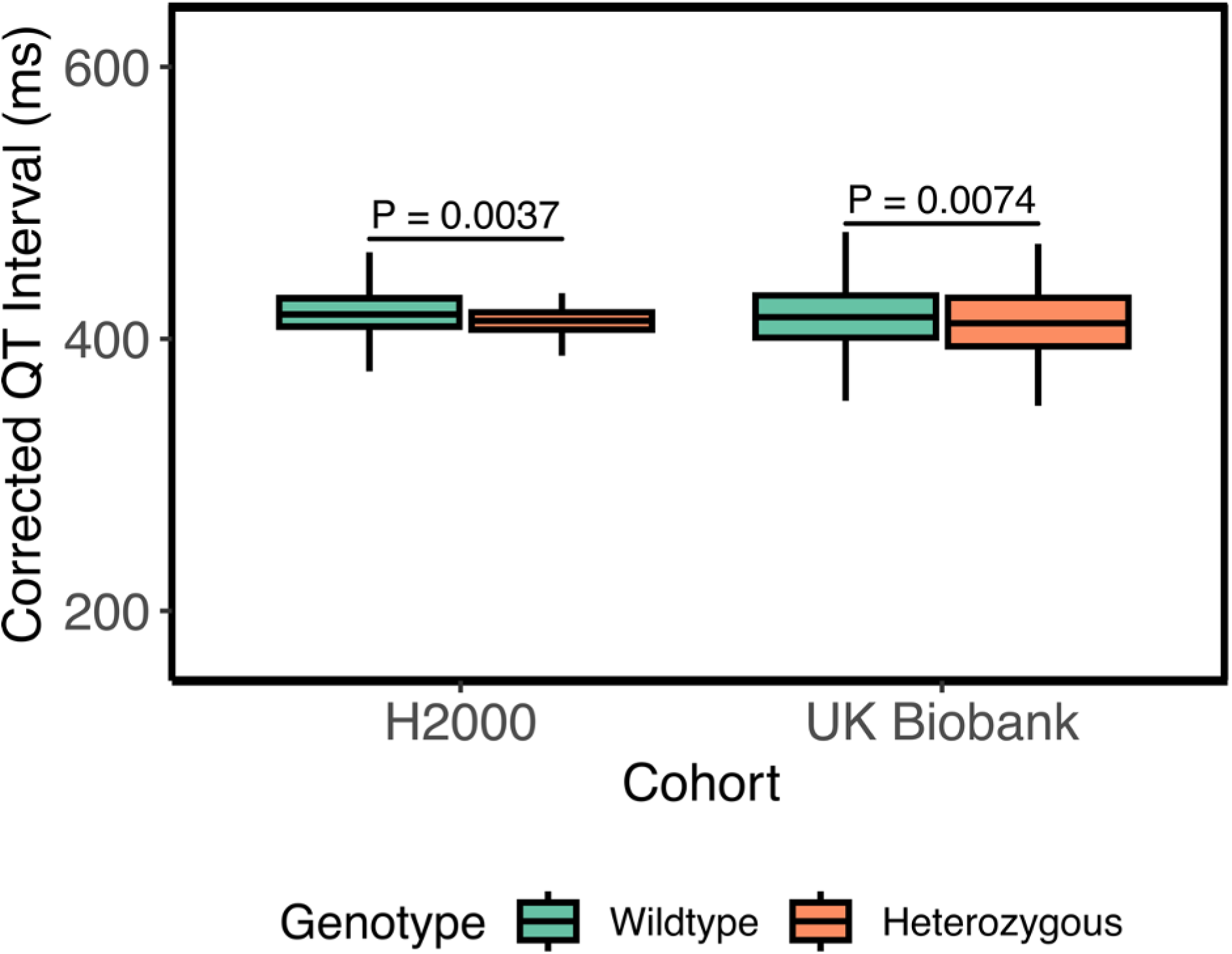
Corrected QT intervals for heterozygotes and wild types of T220I. Data from the H2000 study and the full UKBB dataset (12-lead resting or 3-lead upright ECG before exercise test) are shown as boxplots. Box plot showing the median and IQR with whiskers extending to 1.5*IQR, outliers were omitted for clarity. Significantly shortened corrected QT intervals (QTc) were found in both cohorts, p = 0.0037 for H2000 and p = 0.0074 for the UKBB.

**Figure 5.**
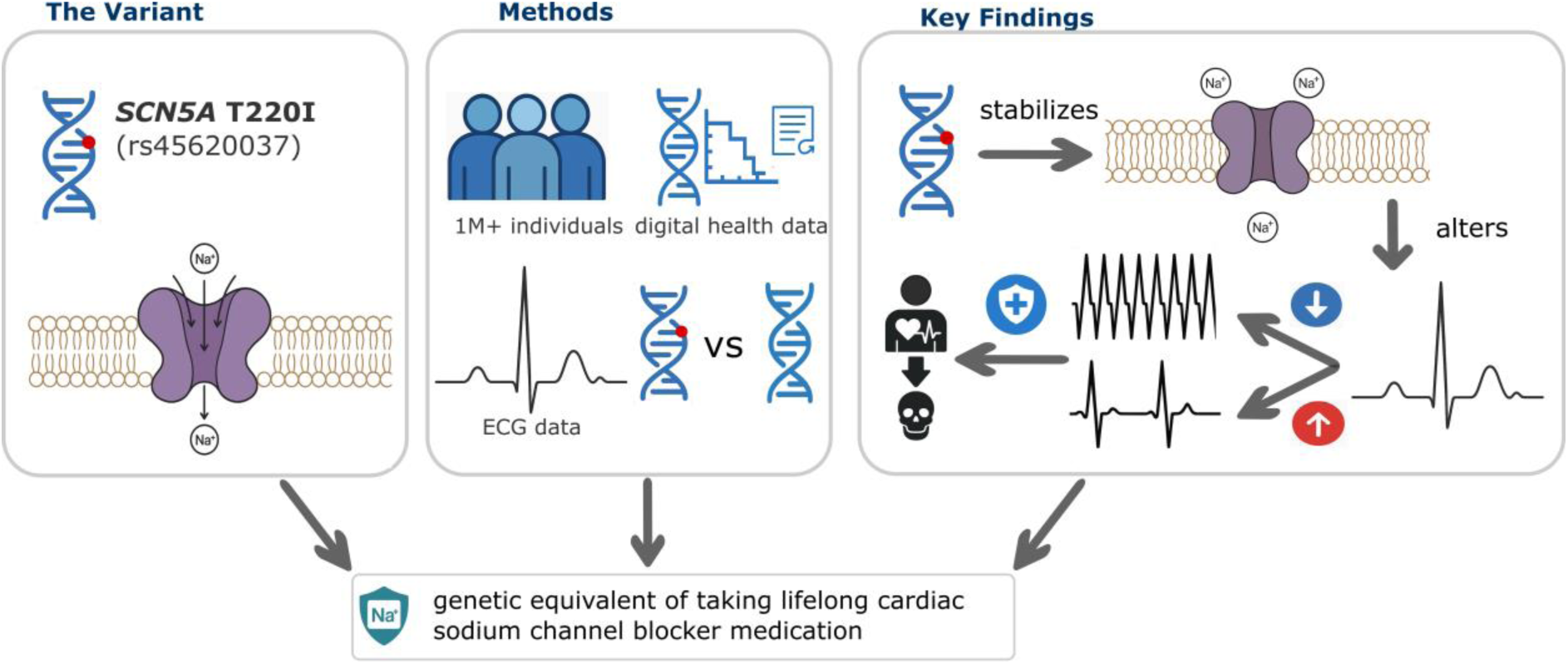
T220I acts as the genetic equivalent of a lifelong cardiac sodium channel blockade. We used genetic data along with electronic health record data and ECG data to show a substantial protection of T220I against tachyarrhythmias with the increased risk for bradyarrhythmias and overall protection against cardiac arrhythmia related death.

## Discussion

The Finnish-enriched *SCN5A* variant T220I is unique in its relatively large protective effect on AFib, with T220I carriers having half the risk of wild types. To the best of our knowledge, no similarly sized protective effects from cardiac arrhythmias or heart failure have been described for any other variants in *SCN5A* or in other genes. Here, we characterized T220I’s effect on cardiac phenotypes and conduction parameters in more than 1M individuals. In agreement with the experimental evidence of a mild LoF effect, our results are consistent with the variant acting similarly to a sodium channel blocker. While protecting against multiple cardiac arrhythmias such as AFib and types of heart failure, T220I also caused typical sodium channel blocker associated ‘side effects’, including increased risk for bradycardia and slowed conduction and specific ECG changes. Overall, the protective effects predominated, resulting in an overall protection against death due to cardiac arrhythmias. We further showed that the variant can effectively reduce the genetic liability for AFib conferred by common variants from a high to an average risk.

### Our data mostly confirms previous associations of T220I with cardiac arrhythmias

The T220I variant has been previously implicated in different cardiac diseases, based on studies of individual families ^11,16,18,24^(for an overview, see also Supplementary Table 1). Here, we critically evaluated these previous findings in a large and uniformly phenotyped cohort. We could confirm our earlier observation of T220I’s protection from AFib ^13^ with an almost tripled cohort size. The effect of T220I is much larger than commonly observed for GWAS variants with carriers having about half the risk of AFib compared to population average.

Previously, T220I has been observed in individuals with childhood-onset SSS of one family, but only together with a larger LoF effect variant on the other allele, in compound heterozygous state. Here, we observed a dosage effect, with an elevated risk for SSS in T220I homozygotes. However, as SSS is most prevalent in older individuals, we could only observe a risk increase in adult age groups, > ca. 50 years for homozygotes and > ca. 65 years for heterozygotes. Due to its incomplete penetrance, the variant should thus be classified as a risk factor ^31^ for adult-onset SSS. Given previous case reports, it thus makes sense that it could also act as a hypomorphic variant contributing to a more severe childhood-onset SSS when in a compound heterozygous state with a LoF variant in agreement with its experimentally found mild LoF effect ^11,17^, while we did not provide direct evidence in this study. In our data, we cannot confirm previous case reports stating T220I may increase risk for AFib or dilated cardiomyopathy, and rather provide evidence of protective effects on these outcomes. Given that the earlier studies were confined to individual families, we find it much more likely that these were chance observations while we were actually powered to investigate such disease associations (see also power analyses, results).

### Protection and risk increases for specific arrhythmias implies both similarities and differences between T220I and known effects of specific sodium channel blockers

Given T220I’s mild LoF effect ^17,18^, we expect that it may act similarly to a lifelong sodium channel blocker. Sodium channel blockers are a class of drugs (Vaughn Williams Class 1 antiarrhythmics) that are often prescribed to control atrial and ventricular cardiac arrhythmias. Their most common side effect is bradyarrhythmia ^32^, slowing of the heart rate via the sinoatrial node, as well as cardiac conduction delays or blocks. This aligns with our observations of an increased rate of SSS, AV block and bifascicular blocks in variant carriers in homozygotes and partially or to a lesser degree in heterozygotes. These findings are also consistent with hypomorphic *SCN5A* variants associated with conduction disorders ^5,6,16^. Other known side effects of class I antiarrhythmic sodium channel blockers are atrial flutter, ventricular tachycardia, ventricular fibrillation, Torsade de pointes or Brugada-like patterns. As multiple of these conditions such as ventricular fibrillation can be directly life-threatening, such side effects have made the indication to prescribe sodium channel blockers for AFib narrow. We were surprised to find no increased risk for these conditions in carriers of T220I even though we were adequately powered to find at least large effects. On the contrary, we found protective effects of T220I from ventricular tachycardia. Different mechanisms could explain the absence of common sodium channel blockers’ side effects. One reason may lie in T220I’s highly specific effect on cardiac sodium channels while common sodium channel blocker drugs, such as quinidine, have more unspecific effects such as additional blockage of potassium channels ^33^. However, increased specificity cannot be the only explanation when taking into account that BrS caused by *SCN5A* LoF and consequently lower Na_V_1.5 expression levels are associated with a highly elevated risk (up to 30%) of ventricular fibrillation or ventricular tachycardia ^34^. Thus, it could be explained by a dosage effect with Na_V_1.5 blockade conferring protection from AFib while not causing ventricular tachycardia within a certain therapeutic window. It seems that T220I may have such protective effects also in a homozygous state within that therapeutic window as we did not observe any case of VF in 28 T220I homozygotes. In addition, we and others ^24^ did not observe any Brugada-like ECG patterns in T220I carriers. Another explanation could lie in the specific mechanism of the variant. From electrophysiology experiments T220I is thought to stabilize the inactivated state of the cardiac sodium channel through a gating pore current ^27^. Due to its location in the voltage-sensor of the channel which is critical for the channel’s response to voltage changes ^27^, T220I was previously thought to open an abnormal Na+ permeation pathway (also called “gating pore”). The altered channel morphology should cause a leakage of Na+ ions during the resting phase of the channel, allowing Na+ to flow into the cardiac cells during diastole of the heart ^27^. This may cause a partial depolarization of the hyperpolarized resting membrane potential stabilizing the inactivated state of the sodium channels. As a result, fewer sodium channels are thought to be available during action potential firing ^26,27^, thus reducing the peak sodium current and slowing the conduction velocity ^17,18^. This could have beneficial effects reducing late sodium current, in line with our observations of shortened QT intervals in T220I carriers. Regarding the effects on QT time, T220I resembles the effects of the weaker class 1B sodium channel blockers such as Lidocaine which are in fact used to treat ventricular tachycardia in emergency situations. Similarly, to lidocaine, T220I also lacks an effect on QRS and P-wave duration. However, a possible prolonged PR interval in T220I carriers is not a known typical effect of Lidocaine ^33^. In addition, the drug is not known to confer protection from chronic AFib, heart failure or DCM. The molecular mechanisms thus seem somewhat different between class 1B sodium channel blockers and T220I. Precise mechanisms of T220I can only be answered by more detailed experimental investigations.

In addition to protection from cardiac arrhythmias, we found protective effects of T220I from dilated cardiomyopathy and (left) heart failure (with nominal significance individually, but consistently in the same direction). This is particularly interesting, as medical rhythm control in AFib is generally thought to not improve outcomes over rate control potentially due to toxic side effects of antiarrhythmic drugs ^35,36^. However, more recent evidence suggests a potential positive effect of rhythm control in early AFib ^37^, a potential paradigm shift in the field^37^ in line with our observations. While AFib prevention may play a role in protecting T220I variant carriers from heart failure, other mechanisms—such as T220I’s protection against premature ventricular depolarization—might also contribute. However, our study was not sufficiently powered to investigate this.

T220I gives us a unique opportunity to investigate the potential risks and benefits of sodium channel blockade in specific scenarios. Following the surprising finding of elevated mortality due to cardiac arrhythmias in patients who received sodium channel blockers after a myocardial infarction in the CAST trial ^30^, sodium channel blockers have generally been considered to be contraindicated in individuals with structural heart disease during the last 30 years (myocardial infarction or heart failure). While we also observed an elevated all-cause mortality in patients with “genetic sodium channel blockade” (T220I carriers) after myocardial infarction, T220I associated mortality continuously decreased 10-15 years post myocardial infarction. This is a promising starting point to motivate evaluation of non-class Ic sodium channel blockers in specific patient populations, in whom new clinical trials would otherwise be considered too risky or unethical. Lastly, in our data, T220I could counterbalance a high disease-specific PGS for AFib, capturing genetic liability for AFib conferred by common variants. Specifically, individuals with high AFib-PGS who carried T220I had only an average lifetime risk of AFib. As such, T220I carrier status may represent an important modifier in personalized prediction models for AFib.

### Limitations

Our study has several limitations. First, while 28 homozygotes are a sizable number for rare variant studies, it limits the power to detect effects on more rare phenotypes. However, given the dosage effects we observe for well-powered phenotypes, we assume extrapolating effects from heterozygotes to be a good approximation. Second, our clinical information in FinnGen is restricted to ICD codes, as we did not have access to detailed clinical notes.

## Conclusion

In this study we found that the genetic *SCN5A* T220I variant acts like a lifelong sodium channel blocker, mostly protecting from diverse cardiac arrhythmias, while also having some typical sodium channel blocker ‘side effects’. Overall, T220I had a net protective effect on mortality due to cardiac arrhythmia. We could use this genetic proxy to learn about potential applications of pharmacological sodium channel blockade. For example, we find that T220I also protects from heart failure contributing to an ongoing debate in the field. We can further investigate sodium channel blockade after myocardial infarction (CAST trial) adding further information that would be unethical to investigate with clinical studies. Experimental workup will be crucial to fully understand the precise effects of T220I and the related therapeutic potential.

## Data Availability

Based on National and European regulations (GDPR) access to individual-level sensitive health data must be approved by national authorities for specific research projects and for specifically listed and approved researchers. The FinnGen data may be accessed through Finnish Biobanks? FinBB portal (www.finbb.fi). For access to the UK Biobank data, the procedures are described at https://www.ukbiobank.ac.uk/enable-your-research

https://www.ukbiobank.ac.uk/enable-your-research

## Acknowledgements

We want to thank Carl Jannes Neuse for carefully reading and commenting on the manuscript. We want to thank Andrea Eoli for his comments about the visualization of figures.

We want to acknowledge the participants and investigators of the FinnGen study, the UKBB and H2000. The FinnGen project is funded by two grants from Business Finland (HUS 4685/31/2016 and UH 4386/31/2016) and the following industry partners: AbbVie Inc., AstraZeneca UK Ltd, Biogen MA Inc., Bristol Myers Squibb (and Celgene Corporation & Celgene International II Sàrl), Genentech Inc., Merck Sharp & Dohme LCC, Pfizer Inc., GlaxoSmithKline Intellectual Property Development Ltd., Sanofi US Services Inc., Maze Therapeutics Inc., Janssen Biotech Inc, Novartis AG, and Boehringer Ingelheim International GmbH. Following biobanks are acknowledged for delivering biobank samples to FinnGen: Auria Biobank (www.auria.fi/biopankki), THL Biobank (www.thl.fi/biobank), Helsinki Biobank (www.helsinginbiopankki.fi), Biobank Borealis of Northern Finland (https://www.ppshp.fi/Tutkimus-ja-opetus/Biopankki/Pages/Biobank-Borealis-briefly-in-English.aspx), Finnish Clinical Biobank Tampere (www.tays.fi/en-US/Research_and_development/Finnish_Clinical_Biobank_Tampere), Biobank of Eastern Finland (www.ita-suomenbiopankki.fi/en), Central Finland Biobank (www.ksshp.fi/fi-FI/Potilaalle/Biopankki), Finnish Red Cross Blood Service Biobank (www.veripalvelu.fi/verenluovutus/biopankkitoiminta), Terveystalo Biobank (www.terveystalo.com/fi/Yritystietoa/Terveystalo-Biopankki/Biopankki/) and Arctic Biobank (https://www.oulu.fi/en/university/faculties-and-units/faculty-medicine/northern-finland-birth-cohorts-and-arctic-biobank). All Finnish Biobanks are members of BBMRI.fi infrastructure (https://www.bbmri-eric.eu/national-nodes/finland/).

## Code availability

Please see https://finngen.gitbook.io/documentation/ for a detailed description of data production and analysis, including the code that was used to run analyses, and https://github.com/FINNGEN/ for further code repositories that were used to run analyses in FinnGen. R code to reproduce figures is available at https://github.com/Waseju/SCN5A.

## Data availability

Based on National and European regulations (GDPR) access to individual-level sensitive health data must be approved by national authorities for specific research projects and for specifically listed and approved researchers.

The FinnGen data may be accessed through Finnish Biobanks’ FinBB portal (www.finbb.fi). For access to the UK Biobank data, the procedures are described at https://www.ukbiobank.ac.uk/enable-your-research

## Ethics and data approval

This study complies with all relevant ethical regulations. We obtained approval for the FinnGen study protocol from the Ethics Committee of the Hospital District of Helsinki and Uusimaa (approval number: HUS/990/2017). The UK Biobank Study’s ethical approval had been granted by the National Information Governance Board for Health and Social Care and the NHS North West Multicenter Research Ethics Committee.

All participants provided informed consent through electronic signature at the baseline assessment. The data used in this study is available in the UK Biobank database under the application numbers of 77717 and 17488, 17788 was approved by the local Massachusetts General Hospital Institutional Review Board.

Data are available in a public, open access repository (https://www.ukbiobank.ac.uk/).

Uusimaa Ethics provided approval for the Health 2000 survey (No. 407/E3/2000). All survey participants provided written informed consent. Access to the Health 2000 survey dataset, and linked hospitalization and death records were granted by the Statistics Finland and National Institute for Health and Welfare (THL) in Finland.

Study subjects in FinnGen provided informed consent for biobank research, based on the Finnish Biobank Act. Alternatively, separate research cohorts, collected prior the Finnish Biobank Act came into effect (in September 2013) and start of FinnGen (August 2017), were collected based on study-specific consents and later transferred to the Finnish biobanks after approval by Fimea (Finnish Medicines Agency), the National Supervisory Authority for Welfare and Health. Recruitment protocols followed the biobank protocols approved by Fimea. The Coordinating Ethics Committee of the Hospital District of Helsinki and Uusimaa (HUS) statement number for the FinnGen study is Nr HUS/990/2017.

The FinnGen study is approved by Finnish Institute for Health and Welfare (permit numbers: THL/2031/6.02.00/2017, THL/1101/5.05.00/2017, THL/341/6.02.00/2018, THL/2222/6.02.00/2018,

THL/283/6.02.00/2019, THL/1721/5.05.00/2019 and THL/1524/5.05.00/2020), Digital and population data service agency (permit numbers: VRK43431/2017-3, VRK/6909/2018-3, VRK/4415/2019-3), the Social Insurance Institution (permit numbers: KELA 58/522/2017, KELA 131/522/2018, KELA 70/522/2019, KELA 98/522/2019, KELA 134/522/2019, KELA 138/522/2019, KELA 2/522/2020, KELA 16/522/2020), Findata permit numbers THL/2364/14.02/2020, THL/4055/14.06.00/2020, THL/3433/14.06.00/2020, THL/4432/14.06/2020, THL/5189/14.06/2020, THL/5894/14.06.00/2020, THL/6619/14.06.00/2020, THL/209/14.06.00/2021, THL/688/14.06.00/2021, THL/1284/14.06.00/2021, THL/1965/14.06.00/2021, THL/5546/14.02.00/2020, THL/2658/14.06.00/2021, THL/4235/14.06.00/2021, Statistics Finland (permit numbers: TK-53-1041-17 and TK/143/07.03.00/2020 (earlier TK-53-90-20) TK/1735/07.03.00/2021, TK/3112/07.03.00/2021) and Finnish Registry for Kidney Diseases permission/extract from the meeting minutes on 4^th^ July 2019.

The Biobank Access Decisions for FinnGen samples and data utilized in FinnGen Data Freeze 12 include: THL Biobank BB2017_55, BB2017_111, BB2018_19, BB_2018_34, BB_2018_67, BB2018_71, BB2019_7, BB2019_8, BB2019_26, BB2020_1, BB2021_65, Finnish Red Cross Blood Service Biobank 7.12.2017, Helsinki Biobank HUS/359/2017, HUS/248/2020, HUS/430/2021 §28, §29, HUS/150/2022 §12, §13, §14, §15, §16, §17, §18, §23, §58, §59, HUS/128/2023 §18, Auria Biobank AB17-5154 and amendment #1 (August 17 2020) and amendments BB_2021-0140, BB_2021-0156 (August 26 2021, Feb 2 2022), BB_2021-0169, BB_2021-0179, BB_2021-0161, AB20-5926 and amendment #1 (April 23 2020) and it’s modifications (Sep 22 2021), BB_2022-0262, BB_2022-0256, Biobank Borealis of Northern Finland_2017_1013, 2021_5010, 2021_5010 Amendment, 2021_5018, 2021_5018 Amendment, 2021_5015, 2021_5015 Amendment, 2021_5015 Amendment_2, 2021_5023, 2021_5023 Amendment, 2021_5023 Amendment_2, 2021_5017, 2021_5017 Amendment, 2022_6001, 2022_6001 Amendment, 2022_6006 Amendment, 2022_6006 Amendment, 2022_6006 Amendment_2, BB22-0067, 2022_0262, 2022_0262 Amendment, Biobank of Eastern Finland 1186/2018 and amendment 22§/2020, 53§/2021, 13§/2022, 14§/2022, 15§/2022, 27§/2022, 28§/2022, 29§/2022, 33§/2022, 35§/2022, 36§/2022, 37§/2022, 39§/2022, 7§/2023, 32§/2023, 33§/2023, 34§/2023, 35§/2023, 36§/2023, 37§/2023, 38§/2023, 39§/2023, 40§/2023, 41§/2023, Finnish Clinical Biobank Tampere MH0004 and amendments (21.02.2020 & 06.10.2020), BB2021-0140 8§/2021, 9§/2021, §9/2022, §10/2022, §12/2022, 13§/2022, §20/2022, §21/2022, §22/2022, §23/2022, 28§/2022, 29§/2022, 30§/2022, 31§/2022, 32§/2022, 38§/2022, 40§/2022, 42§/2022, 1§/2023, Central Finland Biobank 1-2017, BB_2021-0161, BB_2021-0169, BB_2021-0179, BB_2021-0170, BB_2022-0256, BB_2022-0262, BB22-0067, Decision allowing to continue data processing until 31^st^ Aug 2024 for projects: BB_2021-0179, BB22-0067,BB_2022-0262, BB_2021-0170, BB_2021-0164, BB_2021-0161, and BB_2021-0169, and Terveystalo Biobank STB 2018001 and amendment 25^th^ Aug 2020, Finnish Hematological Registry and Clinical Biobank decision 18^th^ June 2021, Arctic biobank P0844: ARC_2021_1001.

---

**Supplementary Figure 1.**
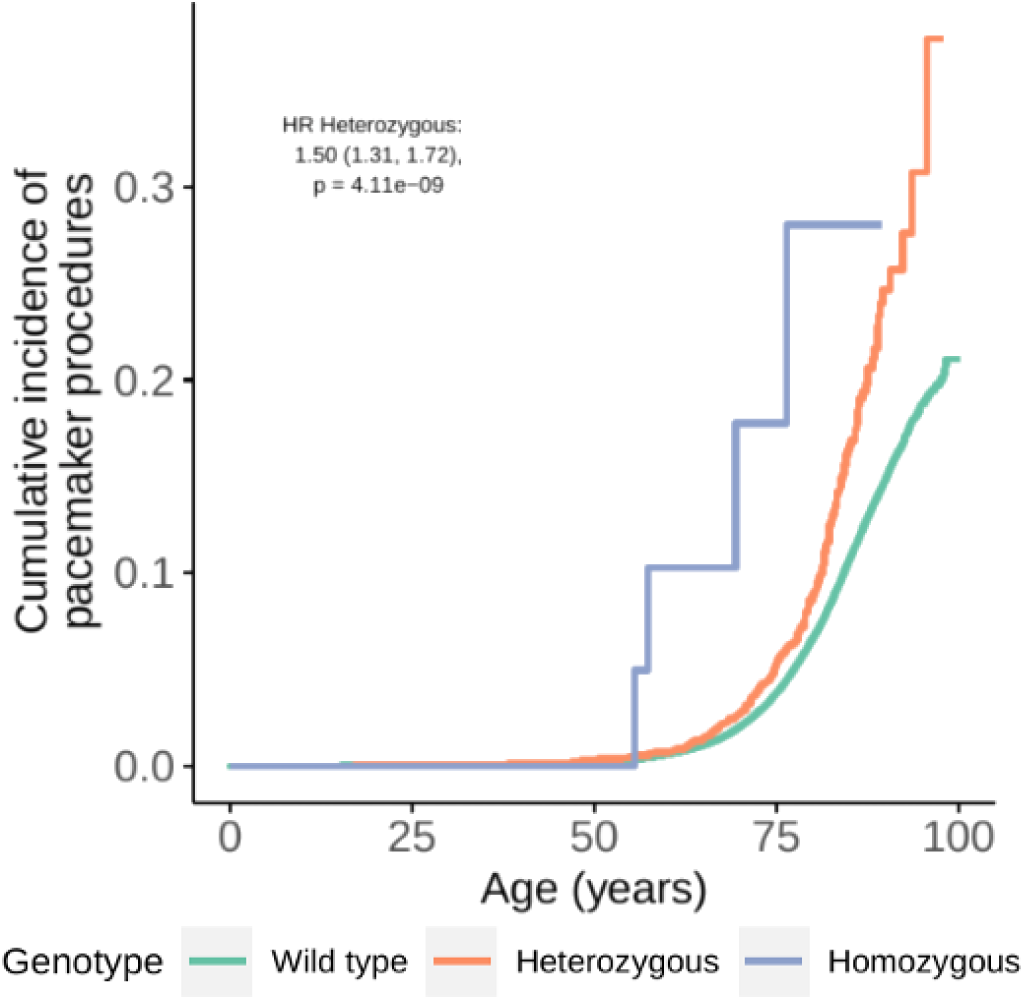
**Age at first pacemaker procedure**, stratified by carrier state of T220I. Data are shown as Kaplan-Meier survival estimates, significant hazard ratios across lifetime are shown inside the survival plots.

**Supplementary Figure 2.**
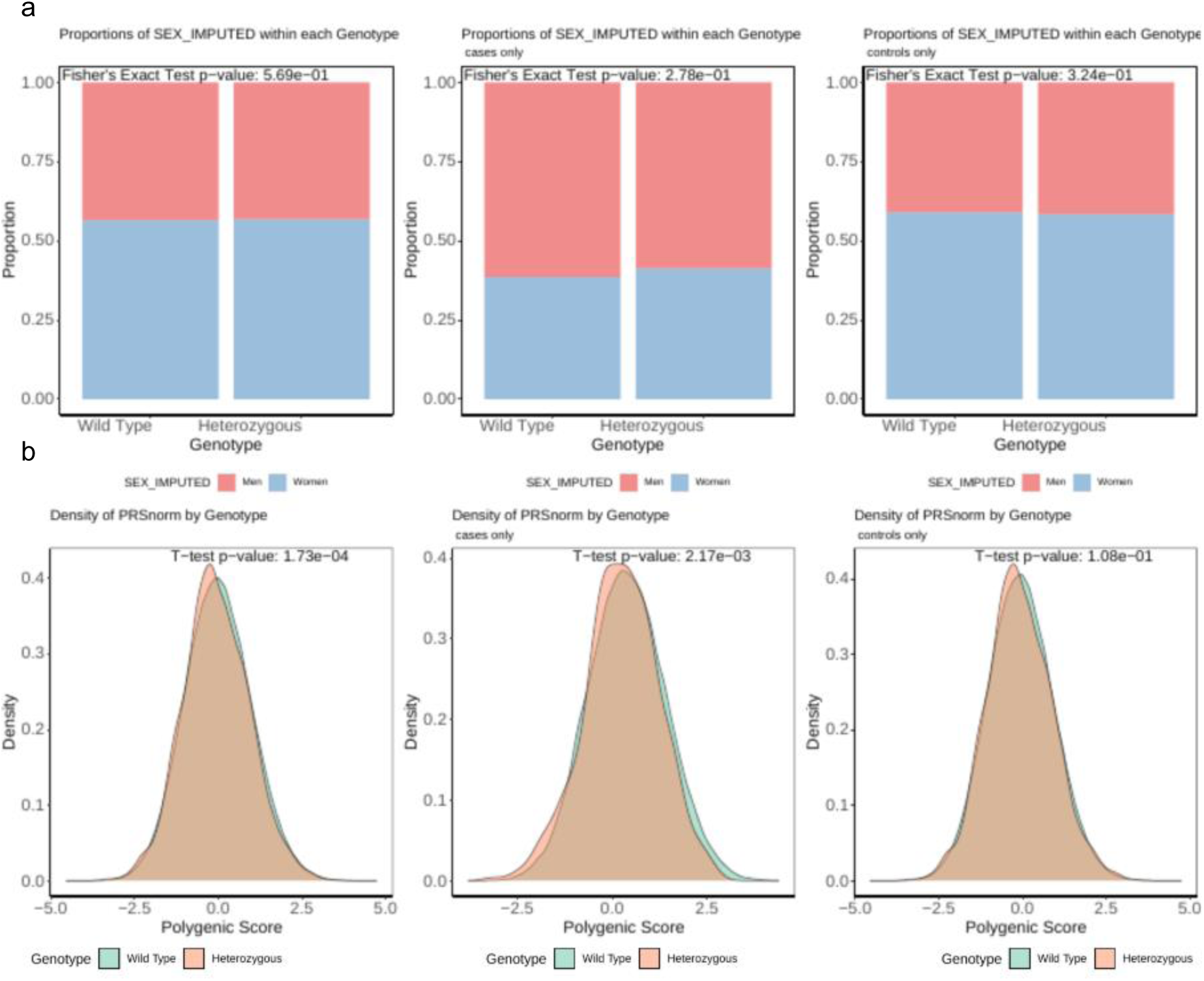
Sex and AFib PGS distributions for T220 Heterozygotes and Wild types in FinnGen DF12. (a) Proportion of Females and Males for all individuals, cases of AFib and controls of AFib. With fisher’s exact test, there are no significant differences for the distribution of sexes between heterozygotes and wild types (p = 0.569). (b) Density plots of an AFib PGS for all individuals, cases of AFib and controls of AFib. With fisher’s exact test, there are significant differences for the distribution of an AFib PGS between heterozygotes and wild types in all individuals (p = 1.73×10-^04^) and cases only (p = 2.17×10^-03^).

**Supplementary Figure 3.**
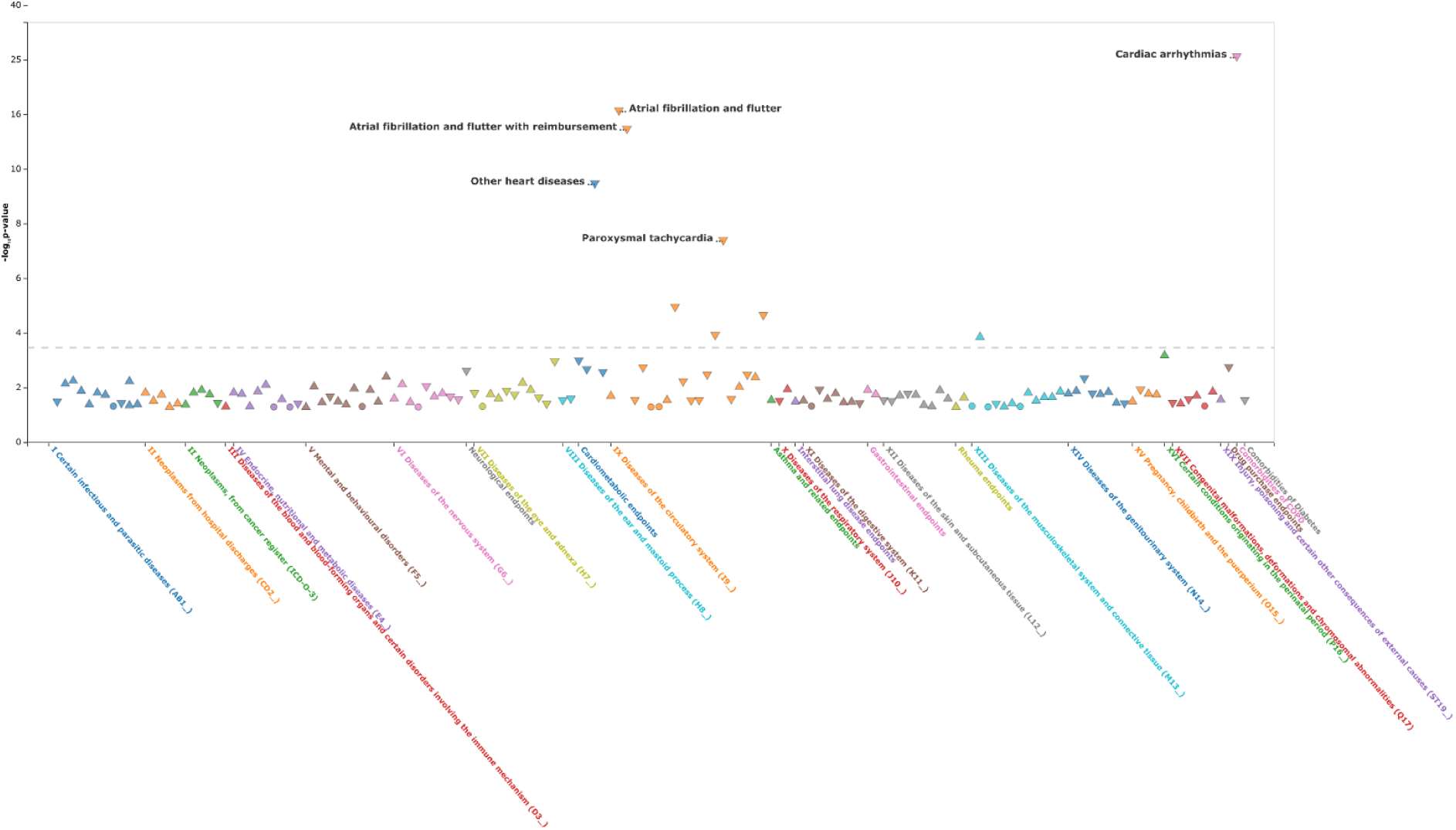
Phenome-wide-association study of T220I. Log10 p-values (y-axis) of associations of the SCN5A variant T220I with all available FinnGen phenotypes (n = 2,469, [x-axis]). The grey dashed line shows the p-values’ multiple testing threshold.

**Supplementary Figure 4.**
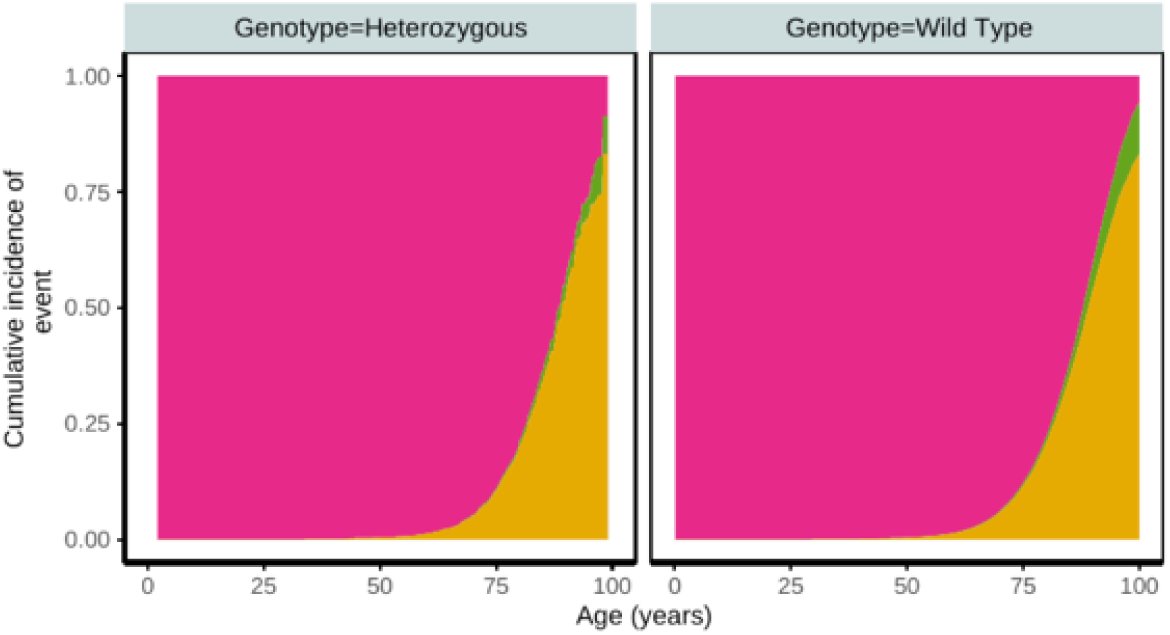
Cumulative incidence of death due to cardiac arrhythmi. **a**, stratified by carrier states of T220I. Panels show cumulative incidence of death due to cardiac arrhythmia (y-axis) across the lifetime (age in years, x-axis) in T220I heterozygotes (left panel) and wild types (right panel), respectively. Heterozygotes showed a decreased risk of death due to cardiac arrhythmias. Here, this is shown with a competing risk analysis, adjusting for other causes of death.

**Supplementary Figure 5.**
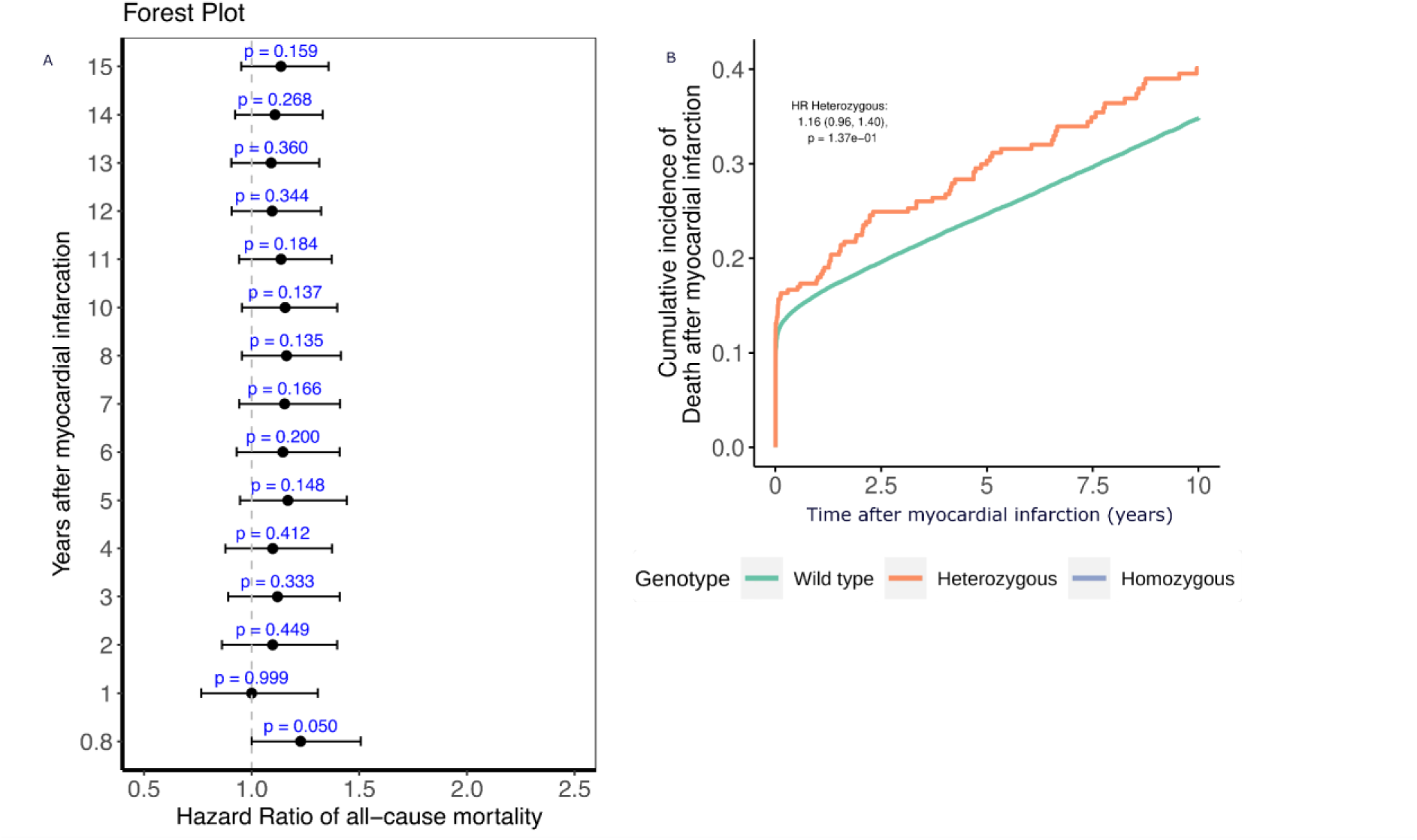
T220I increased all-cause mortality after myocardial infarction. Panel A shows hazard ratios of all-cause mortality post myocardial infarction for T220I carriers, measured in intervals of one year, stratified by carrier states of T220I. Panel B shows cumulative incidence of death (y-axis) in the time (x-axis, years) after a myocardial infarction event Heterozygotes showed an increased risk for all-cause mortality, with stronger effects early after a myocardial infarction.

**Supplementary Table 1.** Summary of studies investigating the SCN5A T220I variant and its association with cardiac arrhythmias.

| Study | Sample Size | Study Design | Main Findings | Cohorts/Database Used |
| --- | --- | --- | --- | --- |
| Heyne et al., 2023 | ~ 190,000 (FinnGen)<br>~ 500,000 (UK Biobank) | Population-based, longitudinal | Protective effect against AFib ($p = 2 \times 10^{-8}$); increased risk for SSS in homozygotes ($p = 9 \times 10^{-4}$); | FinnGen, UK Biobank |
| Benson et al., 2003 | One family (N = 2 affected variant carriers) | Family study | Linked compound heterozygous *SCN5A* mutations, including T220I, to congenital (early-onset) SSS in children. | Individual, family-based genetic data + electrophysiology experiments |
| Gui et al., 2010 | - | Functional study | Demonstrated altered channel kinetics contributing to SSS; identified multiple LoF mechanisms in *SCN5A* mutations, including T220I. | Electrophysiology experiments |
| Olson et al., 2005 | One family (N = 2 variant carriers with DCM, 2 healthy variant carriers with 'larger hearts') | Family study | Association between T220I and DCM; conduction delay of T220I | Selected (DCM) patient populations |
| Olesen et al., 2012 | Case-report (N = 1) | Observational, ECG-focused | Association between T220I and early-onset lone AF; mild LoF effects on Nav1.5 confirmed. | Selected (AF) patient populations |
| Baskar et al., 2014 | Case report (N = 1) | Family study | Reported an 11-year-old girl with atrial standstill carrying compound heterozygous *SCN5A* mutations, including T220I. | Individual family-based genetic data |

**Supplementary Table 2.**
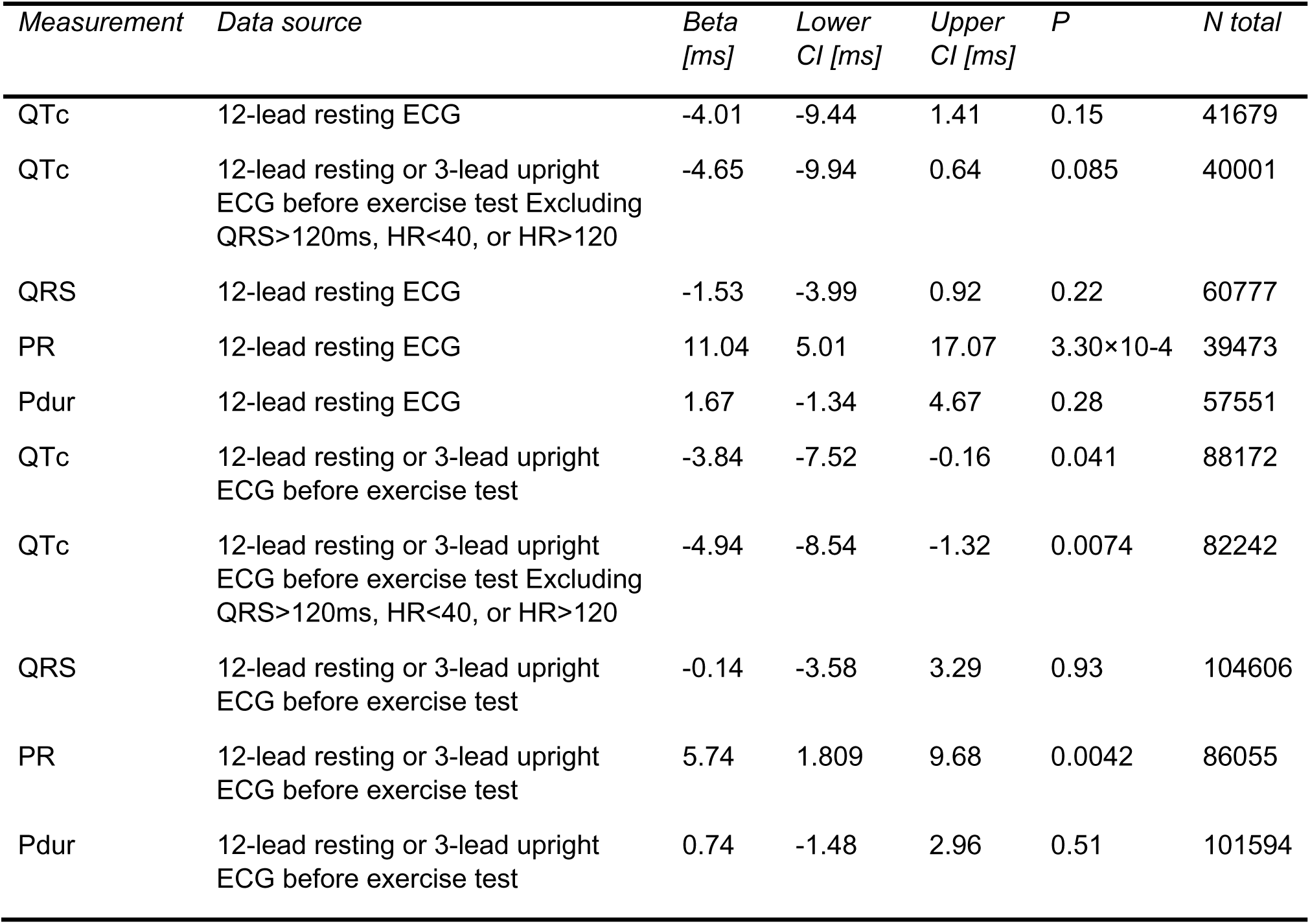
Associations between electrophysiological parameters and genetic variants across different ECG setups.

| *Measurement* | *Data source* | *Beta*<br>*[ms]* | *Lower*<br>*CI [ms]* | *Upper*<br>*CI [ms]* | *P* | *N total* |
| --- | --- | --- | --- | --- | --- | --- |
| QTc | 12-lead resting ECG | -4.01 | -9.44 | 1.41 | 0.15 | 41679 |
| QTc | 12-lead resting or 3-lead upright ECG before exercise test Excluding QRS>120ms, HR<40, or HR>120 | -4.65 | -9.94 | 0.64 | 0.085 | 40001 |
| QRS | 12-lead resting ECG | -1.53 | -3.99 | 0.92 | 0.22 | 60777 |
| PR | 12-lead resting ECG | 11.04 | 5.01 | 17.07 | 3.30×10<sup>-4</sup> | 39473 |
| Pdur | 12-lead resting ECG | 1.67 | -1.34 | 4.67 | 0.28 | 57551 |
| QTc | 12-lead resting or 3-lead upright ECG before exercise test | -3.84 | -7.52 | -0.16 | 0.041 | 88172 |
| QTc | 12-lead resting or 3-lead upright ECG before exercise test Excluding QRS>120ms, HR<40, or HR>120 | -4.94 | -8.54 | -1.32 | 0.0074 | 82242 |
| QRS | 12-lead resting or 3-lead upright ECG before exercise test | -0.14 | -3.58 | 3.29 | 0.93 | 104606 |
| PR | 12-lead resting or 3-lead upright ECG before exercise test | 5.74 | 1.809 | 9.68 | 0.0042 | 86055 |
| Pdur | 12-lead resting or 3-lead upright ECG before exercise test | 0.74 | -1.48 | 2.96 | 0.51 | 101594 |

## Supplement

Supplementary Note 1

In the UK Biobank (n = 487,702), we replicated our PheWAS for T220I in ICD-10 subcategories (I40.0 − I50.9) we found nominally significant protective effects with similar ORs as in FinnGen for paroxysmal atrial fibrillation (OR = 0.44, 95% CI: 0.20-0.99, p = 0.046) and unspecified atrial fibrillation (OR = 0.6, 95% CI: 0.41-0.89, p = 0.01). For broader ICD categories (I40 - I50) we found a protective effect in carriers of T220I for AFib (OR = 0.6, 95% CI: 0.41-0.89, p = 7.6×10^-4^). However, for sick sinus syndrome we found similar, non-significant effects, as in FinnGen (OR = 1.88, 95% CI: 4.56-0.78, p = 0.16).

Supplementary Note 2

To understand how medications may influence the effects of T220I on the incidence of AFib in our results we adjusted a cox proportional hazard model to include prior medication usage, with that our results remained stable (HR = 0.59, 0.53, 0.65, p = 6.62×10^-21^). Further we used a model (Method: generalized linear model) to test our hypothesis that T220I acts independent of prior medication usage, revealing no significant differences (log(OR) = −0.53,SD = 0.74,p = 7.13×10^-13^ and log(OR) = − 0.51,SD = 0.74,p = 7.46×10^-12^) when adjusting for additional medication information.

Supplementary Note 3

In H2000, the association between QTc time remained significant after adjustment for other covariates (calcium level, TSH, diastolic blood pressure, systolic blood pressure, respiratory diseases, arrhythmia, other heart illnesses and first 4 PCs of ancestry), with an average 7.84 ± 2.55 ms, p = 0.0021 decrease in QTcB duration for variant carriers. Notably, we found systolic blood pressure (p = 2.03×10^-12^) and unspecified heart disease (p = 0.014), to influence QTcB time. We found the results also robust to using an alternative QT correction method (−5.83 ± 2.64 ms, p = 0.0027, Method: Framingham correction).

Supplementary Note 4

We found that T220I increased all-cause mortality in the 8 months after a myocardial infarction event (see also Supplementary Figure 5), which is in agreement with the effects of pharmacologic sodium channel blockade after myocardial infarction tested in the Cardiac Arrhythmia Suppression trial (CAST, Echt *et al* NEJM, 1991). In the CAST trial mortality associated with class 1c sodium channel blocker usage was driven by cardiac arrhythmia related mortality. We observed however rather the opposite trend in our data as death due to cardiac arrhythmia was continuously lower when restricting to immediate, underlying or contributing causes of death. Considering other common causes of death after myocardial infarction, we did not see increased rates for mortality due to cardiovascular disease, nor heart failure but rather other causes of death.

As an additional sensitivity analysis, we compared causes of deaths of different causal categories (underlying, immediate and contributing) after MI. We found no significant effects of T220I on death due to AFib, heart failure or cardiovascular causes excluding heart failure and AFib when comparing the different types of cause of death. However, we cannot conclude if this is due to a low sample size or lack of effect. However, we found that the effect on all-cause mortality after MI is mostly driven by contributing and immediate causes of death of other diseases such as chronic obstructive pulmonary disease or other pneumological diseases.

